# Smartphone-based behavioral profiling for distinguishing Dementia with Lewy bodies from Alzheimer’s Disease

**DOI:** 10.1101/2025.03.31.25324871

**Authors:** Gajanan S. Revankar, Abhishek C. Salian, Tatsuhiko Ozono, Maki Suzuki, Hideki Kanemoto, Kazue Shigenobu, Yoshiyuki Nishio, Kenji Yoshiyama, Yuki Yamamoto, Seema S. Revankar, Akihiro Watari, Kota Furuya, Issei Ogasawara, Natsuki Yoshida, Chizu Saeki, Yuta Kajiyama, Daisaku Nakatani, Mamoru Hashimoto, Yoshitaka Nagai, Manabu Ikeda, Etsuro Mori, Ken Nakata, Hideki Mochizuki

**Affiliations:** Center for Global Health, Department of Medical Innovation, Osaka University Hospital, Osaka, Japan; Department of Neurology, Graduate School of Medicine, Osaka University, Osaka, Japan; Thought Petal Technologies, Dandeli, India; Department of Behavioral Neurology and Neuropsychiatry, United Graduate School of Child Development, Osaka University, Osaka, Japan; Department of Psychiatry, Graduate School of Medicine, Osaka University, Osaka, Japan; Department of Medical Innovation, Osaka University Hospital, Osaka, Japan; Department of Health and Sport Sciences, Graduate School of Medicine, Osaka University, Osaka, Japan; Department of Neurology, Kawasaki Medical School, Okayama, Japan; Department of Psychiatry, Kindai University Faculty of Medicine, Osaka, Japan; Department of Neurology, Kindai University Faculty of Medicine, Osaka, Japan; Health and Counseling Center, Osaka University, Osaka, Japan

**Author notes:** Correspondence: Gajanan S. Revankar Center for Global Health, Dept. of Medical Innovation, Graduate School of Medicine, Osaka University Hospital 2-1, Yamadaoka, Suita, Osaka, Japan – 5650871.

## Abstract

**Background and Objective:** Dementia with Lewy bodies (DLB) is frequently misdiagnosed as Alzheimer’s disease (AD) due to overlapping clinical presentations. In this study, we evaluated a smartphone-based digital platform that leverages behavioral markers to accurately differentiate between DLB and AD.

**Methods:** We conducted a cross-sectional study on 81 participants (Healthy controls = 40, AD = 21, DLB = 20), administering MMSE, and a smartphone-based noise pareidolia test (NPT) with integrated eye-tracking and speech inputs. Ground truth for patients were established using nuclear imaging (DAT-SPECT, IMP-SPECT, and/or MIBG). Core behavioral features such as response efficiency, gaze parameters (fixation count, saccadic patterns, surface area coverage) and acoustic features (latency, spectral properties) were extracted. A two-tiered classification system involving deep learning and support vector machine model was used. Based on the behavioral output, probability scores were generated with a focus on explainability to differentiate DLB from AD.

**Results and interpretation:** DLB group showed more pareidolias than AD group on the digital app. DLB patients exhibited several short saccades, inconsistent scanning behavior and smaller fixation dispersion, suggesting impaired top-down modulation of visual attention. However, fixation duration did not differ between AD and DLB. Vocal responses (latency, spectral properties) lacked specificity for DLB classification. Response fluctuation analysis revealed lower variability in AD patients compared to DLB. The app outperformed paper-based methods with 87% sensitivity and 93% specificity, and overall accuracy of 90% for AD–DLB classification. Two AD patients were classified as healthy controls (probability scores: 15%, 45%), likely due to early disease stage or unmeasured neurocognitive factors.

**Conclusions:** By effectively capturing core symptoms, our smartphone-based modality provides a reliable, scalable, and non-invasive tool for classification of DLB from AD. The integration of machine learning enabled precise quantification of behavioral microfluctuations between AD and DLB subtypes, underscoring its significant diagnostic potential. Future research is needed in refining classifiers to address early-stage cases and for disease staging.

## Introduction

Dementia with Lewy bodies (DLB) is the second most common neurodegenerative dementia after Alzheimer’s disease (AD), and together, they make up approximately 75% of all neurodegenerative dementia cases^1,2^. Misdiagnosis or under-diagnosis is common in DLB owing to substantial clinical and pathological overlaps with AD, delaying diagnosis and management in such patients^3–5^. The implication of accurate diagnosis is immediate, since DLB, compared to AD, progresses more quickly, and places a heavy burden on caregivers with no effective disease modifying treatment^6–9^.

Nuclear imaging techniques such as ^123^I-2β-Carbomethoxy-3β-(4-iodophenyl)-*N*-(3-fluoropropyl) nortropane (^123^I-FP-CIT) dopamine transporter single photon emission computed tomography (DAT-SPECT), ^123^I-IMP (Iodine-123 Iodoamphetamine) perfusion SPECT (IMP-SPECT) and ^123^I-metaiodobenzylguanidine (MIBG) myocardial scintigraphy among the gold-standard methods for confirming the diagnosis of DLB^10,11^. As indicative biomarkers, IMP-SPECT, DAT-SPECT, and MIBG scintigraphy demonstrate approx. 80% to 88% sensitivity and 85% to 96% specificity in differentiating DLB from AD, though these values may vary across studies depending on patient populations, imaging protocols, and diagnostic criteria^10,12–14^. While these tools play a crucial role in clinical decision-making for treatment and management, they are highly resource-intensive, with significant costs associated with the personnel and infrastructure required to maintain such scanners^15^. Besides accessibility challenges in non-urban settings, patients with cognitive decline often have compliance issues due to claustrophobia, anxiety or agitation associated with radiological imaging^16,17^.

Nuclear imaging biomarkers directly quantify perfusion levels or dopaminergic degeneration in the brain, whereas behavioral biomarkers from paper tests or digital assessments reflect clinical or behavioral outcomes. However, their limited precision and reliability restrict their use to screening, prompting interest in enhancing these assessments for better detection of disease-specific dysfunctions. For example, the pareidolia test, a neuropsychological tool, leverages visuo-perceptual phenomena to detect and quantify illusions in neurodegenerative diseases^18^. In AD^19,20^, DLB^21,22^ or Parkinson’s disease (PD)^23,24^, pareidolia serve as early behavioral markers for addressing non-cognitive symptoms establishing it as a significant prognostic indicator for disease progression^22^. However, individual variations of pareidolia is common among cognitively normal adults and their utility in differentiating AD and DLB is limited due to low sensitivity^25,26^.

Recent advancements in machine learning and data analysis have significantly bridged the gap between the limitations of traditional behavioral biomarkers and the need for more precise, reliable diagnostic tools^27–29^. These technological innovations enable digital versions of standardized tests to capture and analyze complex microfluctuations in behavior that often elude standard observational methods. Various modalities, including gaze-tracking, speech analysis, motor assessment, and other behavioral variables, have been successfully integrated into digital platforms to extract disease-specific signatures^30–33^. By leveraging machine learning and computational modeling, these digital tools enhance the precision, reliability, and scalability of behavioral biomarkers, allowing for a more precise mapping of downstream behavioral function. In the context of AD and DLB, where early detection remains a critical challenge, such advancements hold transformative potential.

To that end, we developed a smartphone-based application with in-house developed eye-tracking and speech-processing algorithms on a uniquely designed testing paradigm that incorporates the pareidolia test^34,35^. With respect to our study population, the app specifically targeted behavioral signatures tied to three core DLB symptoms^10^ - visual hallucinations, parkinsonism, and fluctuating cognition—to differentiate DLB from AD more effectively than conventional methods. This digital platform was engineered to simultaneously measure visual, motor, and cognitive behavioral signatures, thereby enhancing diagnostic accuracy. Building on our working group’s prior experience on visuo-perceptual testing paradigms^34,36,37^, this effort aimed to enhance the test’s prognostic value while generating a comprehensive behavioral profile that aligns closely with the pathological underpinnings of DLB.

In summary, the aim of this study was to validate the effectiveness of our technology in differentiating DLB from AD, assessing its sensitivity, specificity, and real-world applicability. By comparing behavioral metrics derived from our application to conventional clinical assessments and neuroimaging data, we explored its potential role in enhancing precision medicine approaches for neurodegenerative disorders.

## Methods

### General information

40 Healthy controls (HC), 21 AD, and 20 DLB participants were enrolled in this hospital-based, cross-sectional, prospective study. We employed non-probability sampling, which enabled us to efficiently gather sufficient data within a limited timeframe. AD and DLB patients were recruited from three different hospitals across Osaka, Japan and HCs were sampled from a single center within Osaka University hospital between September 2023 to April 2024. The Osaka University institution review board cleared the protocol for the study to be performed in the Department of Neurology and the Department of Psychiatry, Osaka University, Nippon Life Hospital and Asakayama General Hospital, all located in Osaka, Japan in accordance with the ethical standards of the Declaration of Helsinki (IRB Approval number – 22307). All participants provided written informed consent for the experiment and to publish their clinical data in the study.

For the patient’s cohort, inclusion criteria were (i) ≥ 40 years of age, (ii) diagnosed as AD or DLB according to their clinical diagnostic criteria, (iii) diagnosis of DLB confirmed by at least one positive nuclear imaging biomarker, IMP-SPECT, DAT-SPECT and/or MIBG, (iv) diagnosis of AD ruled out by negative DAT-SPECT and a negative MIBG scan, and (v) able to use a smartphone independently. Patients with any of the following conditions were excluded: (i) if the attending physician/experimenter judged if a patient had severe behavioral, speech or motor impairment hampering the usage of a smartphone, (ii) known eye-related pathologies, (ii) those on antipsychotic medications (olanzapine, risperidone, clozapine, quetiapine), (iii) major psychiatric diagnosis that affected activities of daily living, and (iv) uncontrolled major medical illness such as seizures or cardiovascular diseases.

Healthy participants HC were ≥ 40 years of age, without any past or present neurological problems. Participants were tested with a near-vision Snellen chart integrated in the smartphone to include normal or corrected-to-normal vision subjects only.

Clinical and neuropsychological assessments were conducted by a clinical psychologist or a neurologist, all performed in a single out-patient visit. At the time of testing, all patients had confirmed diagnosis and were receiving either Donepezil and/or Levodopa, along with other non-neurological medications. Most patients with DLB had a history of visual hallucinations, but during the assessment, none of the patients (both AD and DLB) reported any recent complaints of hallucinations.

### Experiment flow

Following informed-consent, participants underwent 3 tests in the following order – i) Paper Noise pareidolia test (NPT)^18^, ii) Mini-mental State Examination, Japanese version (MMSE-J)^38^ and iii) the smartphone app (henceforth mentioned only as ‘app’). Participants performed the app on an Android smartphone [Samsung A32 with screen dimensions 164mm (h) x 76mm (w), portrait mode], that was fixed on a mount, with participants sitting at 20-30 cm from the screen. Although participants were motivated to use the app independently, examiners aided when requested. The app was designed to be completed in approximately 15 minutes and consisted of a calibration module, and a core pareidolia testing module. A brief description of the modules are described below.

#### Calibration module

We positioned participants in a way that avoided creating glares, reflections, or shadows on the testing apparatus. Brightness values adjustments for face were set between 50 – 89 pixels intensity (brightness levels calculated as per RGB pixel values) so that face and eyes were clearly visible. Testing rooms were illuminated via standard multiple fluorescent light sources (luminosity range approx. 250 to 400 lux). For eye-movement calibration, participants were asked to fixate on a pulsating black circular dot stimulus that appeared on a light-grey background with sufficient contrast. A 9-dot calibration sequence was deployed, with the dot, approx. 1cm in diameter, moving slowly from top-left to the bottom-right in a zig-zag fashion for a total time of approx. 30 s. Speech calibration included the users speaking out a predefined sentence that took approx. 7-10s. A digital near-vision Snellen chart was integrated into the smartphone to include normal or corrected-to-normal vision subjects only. The use of eyeglasses did not interfere with testing, as eye movement data was previously shown to remain stable under such conditions^35^.

#### Pareidolia test module

The test comprised of 40 black and white images, with a face embedded in 8 of the 40 images. Participants were to locate and identify faces accurately, and any misidentification of noisy areas as faces were marked as pareidolia. Participants provided speech-based responses in the pareidolia module, answering ‘yes’ or ‘no’ to indicate whether the displayed stimulus contained a face. If they perceived a face, they were also required to tap on its location to confirm their response. The app then automatically advanced to the next screen upon receiving an answer. For onboarding, users completed an interactive practice session where they tested six sample images, both with and without instructions. This approach helped them familiarize themselves with the test objectives and process. Core test images (40) were randomized to prevent a learning effect, although this effect is known to be minimal or non-existent^39^. The validity of the digital pareidolia module against the paper-based pareidolia testing has been demonstrated in our prior work^35,36^.

Figure-1 shows the overall methodology of the study. Legend: IRB = Institutional review board; AD = Alzheimer’s disease; DLB = Dementia with Lewy Body; MMSE-J = Mini mental state examination, Japanese version; NPT = Noise pareidolia test; SVM = Support vector machine model. We screened patients with a clinical diagnosis and included those with a nuclear imaging result. Healthy controls performed only the digital version of the NPT since it was highly correlated to paper-testing^36^. The experiment was conducted in a controlled lab environment, with the smartphone securely mounted at a predetermined distance and angle. Data anonymization protocols involved segregating personal identifiable information (PII) from non-PII data. Using unique identifiers and employing robust encryption techniques, we ensured that sensitive details remain securely isolated from clinical and behavioral datasets, maintaining compliance with Osaka University’s data privacy regulations.

**Figure-1:**
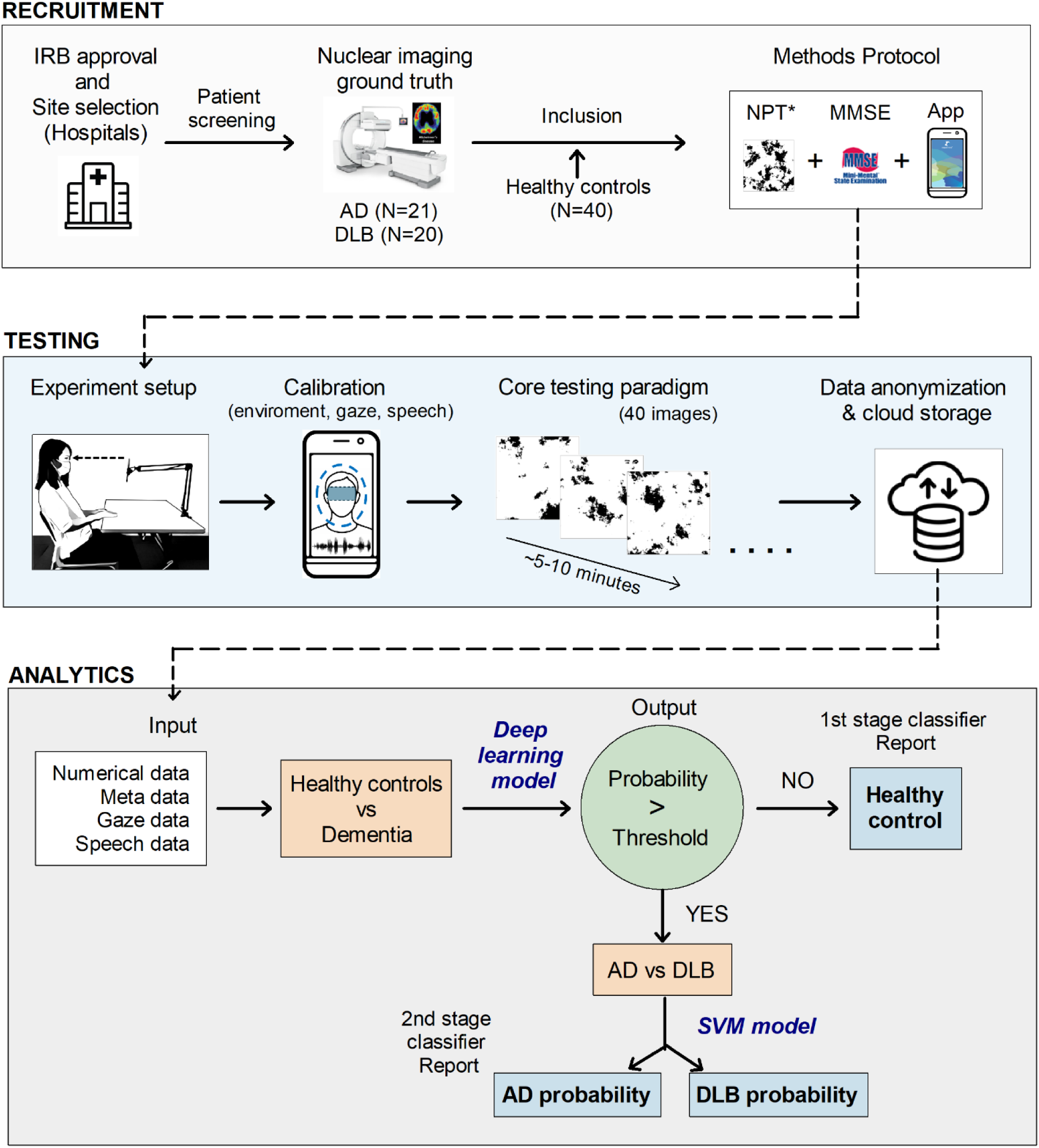
Overview of recruitment, testing and analytical methodology.

### Model details

In our previous study, for eye-tracking, we used a multi-layer feed-forward Convolutional Neural Network (CNN) for base model training^35^. The base CNN model was trained first on MIT GazeCapture dataset^40^ and then fine-tuned on a subset of the healthy elderly Japanese population (N=52 participants). Once the Japanese general model was established, personalization was performed for each new participant. Through our calibration process, a set of data points was rapidly collected, allowing the model to adjust to the individual’s specific features. This personalized model was then used for more accurate gaze prediction for the participants, improving the overall reliability of our system^35^.

#### Training and Test data

We began by merging all data points from both healthy individuals and dementia patients. Each data point was labeled to reflect its corresponding group following which the data was split into a ratio of 80:20. Given the modest overall sample size, relying on a single 80:20 split posed risks of high variance in performance estimates and potential bias toward the specific data partitioning. We therefore implemented k-fold cross-validation – a robust method for performance evaluation with limited data^41^. Specifically, we split the dataset into *k* subsets (folds) with a size of 5 to 10 samples. We then trained the model for 20 iterations, each time using *k*−1 folds for training and the remaining fold for testing. By averaging key performance metrics (such as accuracy, sensitivity, and specificity) across all folds, we obtained a more reliable and comprehensive estimate of the model’s performance. This method reduced the impact of random fluctuations in any single train-test split and provided a better insight into the model’s generalizability.

#### Classifier details

Our classification followed a two-step tiered approach. We first distinguished healthy controls (HC) from those with dementia (AD+DLB) using a custom deep learning model. Here, neural networks (deep learning models) excel at capturing complex patterns in large datasets and can often achieve high accuracy in broad classification tasks (e.g., healthy vs. all dementia types combined). Following a predefined threshold, we then applied Support Vector Machine (SVM) to classify AD from DLB. After the first model narrowed the problem space to dementia cases only, the second step dealt with a smaller subset of data focusing on subtle clinical distinctions between AD and DLB. In this scenario, a simpler model like SVM was effective and more interpretable, since our dataset for second classification was relatively small. Inputs to the algorithm included gaze capture, speech, response accuracies, and stimuli response times.

### Data collection and processing

Each individual provided their explicit and written informed consent to data collection that informed them about collecting the front-facing camera feed for research analyses purposes, and the potential risks involved in performing gaze tasks for several minutes (e.g., eye strain, fatigue). All participants retained the option to have their data deleted at any time. All clinical data were stored in paper-based case report forms. Digital data from the app were saved on a database in a cloud server (AWS) with the processing and readouts done offline.

Data was collected through an android app operating on OS version 13. While the app was functional from OS versions 11 onwards, formal testing on versions older than v13 was not conducted. Analysis of eye-movements were based on the methodological principles as defined in our prior research^34,35^. Briefly, the front-camera video records participants’ gaze for each of 40 stimuli images on the smartphone screen. Images from the 13-megapixel front-facing camera were recorded at 1080p resolution at 30 Hz (frames per second - fps) with timestamps synchronized with the marker location. For the above fps, fixations were defined as any 30 frames of continuous participant gaze within the same region, indicating a single fixation. A gaze cluster threshold of 50 pixels of distance for every continuous pair of gaze predictions was set after a trial-and-error estimation of the optimal count required to plot the visual tracking geometry. Saccades were defined as distances between 2 successive fixations that exceeded this threshold. Together, these metrics were used to calculate a cluster heat-map index i.e. fixation and saccade counts, for comparing the visual tracking for target (faces) and non-target (noise) stimuli.

In the speech module, each participant’s audio data was processed through a standardized pipeline. Audio files in WAV or M4A format were converted to a consistent format (WAV) and trimmed to 30 seconds with a 0.5-second offset. Subsequently, we extracted mel spectrograms from these standardized signals using a dedicated function, which transformed the raw audio into a time-frequency representation suitable for deep learning analysis. These spectrograms were passed through our PyTorch-based model in evaluation mode to obtain output probabilities.

### Statistical analysis

Statistical analysis was performed on JASP (version 0.19.1). For demographic comparisons, Welch’s t-test or Mann Whitney-U test were performed depending on satisfactory assumptions of normality. From both machine learning models, output scores were reported as probabilities. For each of the core test stimulus images (N=40), the app computed the likelihood of the patient’s behavior aligning with either AD or DLB, with the final probability score derived by averaging the values across all stimuli. For these probabilities, we calculated Shapley values which quantify the marginal contribution of each feature to the model’s prediction, providing an interpretable measure of feature importance^42^. To evaluate intra-group variability, we computed the coefficient of variation (CV) for Shapley values within AD and DLB cohorts. The CV was used to assess the consistency of feature importance across patients. Features with high CV indicated greater inter-individual variability, whereas those with lower CV suggest more stable predictive contributions.

For evaluating the effectiveness of a diagnostic test in distinguishing between controls, AD and DLB cases, the Bayesian statistics module provided a framework for analyzing diagnostic performance metrics such as sensitivity, specificity, positive predictive value (PPV), negative predictive value (NPV), and accuracy^43^. We employed this methodology since our dataset was small. A Bayesian receiver operating characteristic (ROC) analysis was used to evaluate the optimum thresholds for sensitivity and specificity of the different modalities to differentiate DLB from AD or controls. Parametric estimates were based on 10,000 MCMC samples, 1000 burn-in samples and 4 runs for convergence.

### Data and Code Availability Statement

The data analyzed in this study will be provided by the corresponding author upon request, subject to reasonable conditions. The paper version of NPT is accessible under an open license for research purposes. The software code utilized in this study relies on internal tooling and infrastructure and is protected by patent (application number: JP2022-179766; RO/JP 52733WO). Gaze estimation was done using PyTorch (v2.1.0) running on Python 3.10. Speech files in WAV format were directly loaded using librosa and for M4A files, conversion to WAV was performed using pydub. Image preprocessing was performed using mediapipe v0.10.0, and gaze maps for the deep learning model were generated using heatmappy library (a Python library).

## Results

### General overview

Participant demographics are summarized in Table-1. Mann-Whitney U-tests for healthy control (HC) and dementia group showed significant differences for age (U = 1259.0, *p = 0.001*), MMSE-J (U = 40.0, *p = 0.001*) and App completion time (U = 1142, *p = 0.002*).

**Table-1:** Demographic data of participants. **Table 1 Legend:** MMSE-J = Mini mental state examination, Japanese version; NPT = Noise pareidolia test; IMP-SPECT – Iodine-123 Iodoamphetamine Single Photon Emission Computed Tomography; DAT-SPECT – Dopamine Transporter Single Photon Emission Computed Tomography; MIBG – Metaiodobenzylguanidine Myocardial Scintigraphy; All scores are presented as means ±standard deviation for descriptive purposes.

|  |  | Healthy controls<br>(N = 40) | Dementia group (N = 41) |  |
| --- | --- | --- | --- | --- |
|  |  |  | Alzheimer's<br>disease<br>(N = 21) | Dementia with<br>Lewy Body<br>(N = 20) |
| Age (years) |  | 67.8 ± 6.9 | 72.6 ± 7.0 | 73.7 ± 5.1 |
| Gender |  | 23F, 17M | 13F, 8M | 4F, 16M |
| MMSE-J |  | 29.0 ± 1.3 | 18.8 ± 5.4 | 20.2 ± 4.7 |
| Disease duration (years) |  | - | 3.1 ± 1.6 | 3.9 ± 1.6 |
| First imaging done<br>(years ago) |  | - | 1.9 ± 1.4 | 2.6 ± 1.4 |
| Noise Pareidolia test (paper)<br>score |  | - | 5.6 ± 6.9 | 8.6 ± 11.3 |
| Imaging<br>scans (N) | SPECT | - | 14 | 6 |
|  | DAT-SPECT | - | 3 | 13 |
|  | MIBG | - | 4 | 14 |
| App core test time (min) |  | 5.5 ± 1.7 | 8.5 ± 2.5 | 7.8 ± 2.9 |

T-tests for AD vs DLB groups did not show significant differences for age, MMSE-J, disease duration or paper Noise Pareidolia Test (NPT) method. However, DLB patients underwent imaging-based diagnosis earlier than AD patients (U = 131, *p = 0.032*). MMSE-J showed a strong negative correlation with NPT for DLB [Spearman’s correlation, r(18) = -0.67, *p = 0.001*] and not for AD [r(19) = -0.29, p = 0.197] (Figure-2).

**Figure-2:**
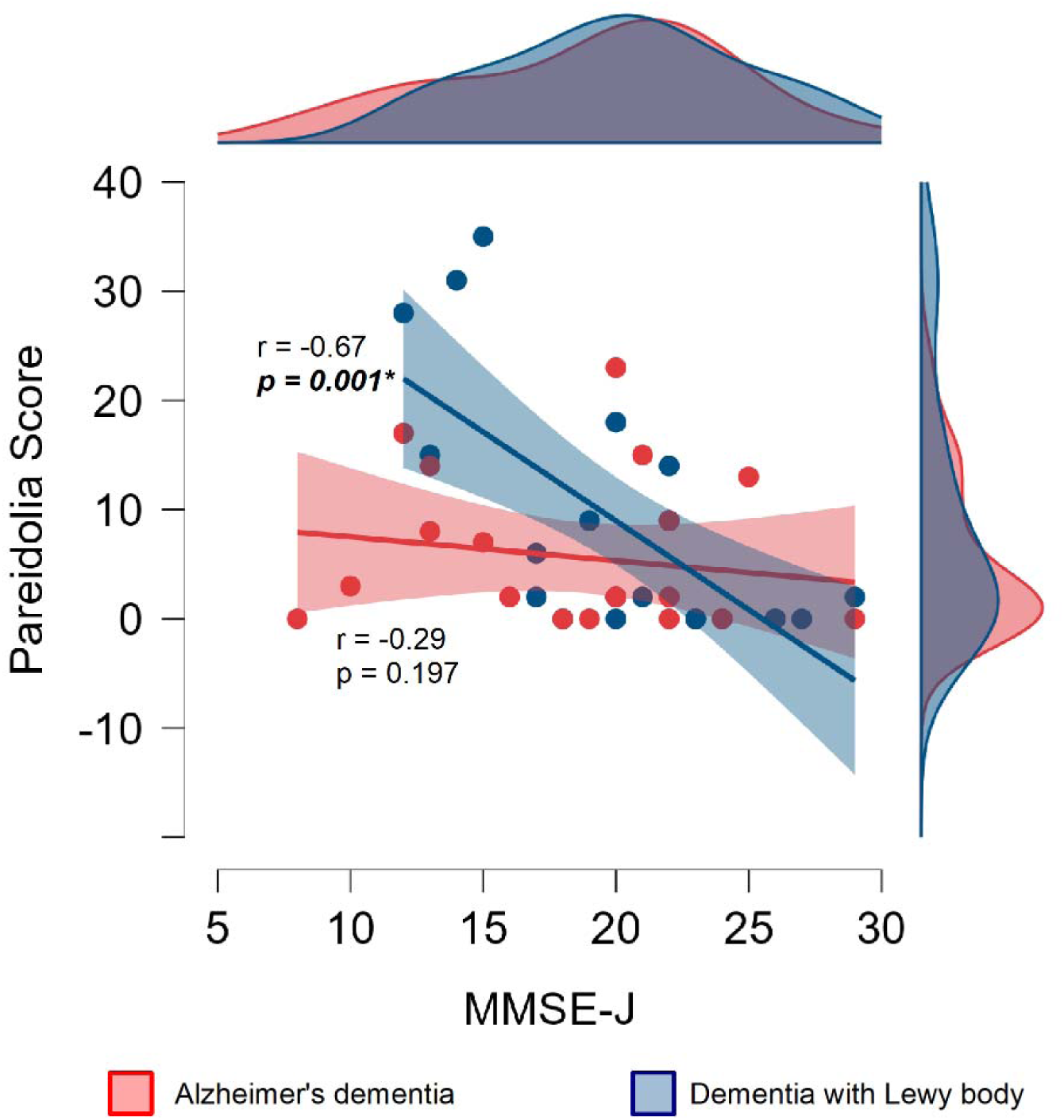
Correlation plot of MMSE-J with pareidolia score in AD and DLB.

Figure-2 shows correlation and distribution (outer axes) of MMSE and Pareidolia scores for AD (red) and DLB (blue) patient groups. DLB patients showed a strong negative correlation, suggesting overlaps in cognitive and perceptual impairments, whereas the tests remained relatively independent domain-wise in AD patients.

Although the healthy control cohort was included for methodological clarity, our primary focus of the results was on the app’s performance in classifying AD and DLB. It is worth noting that, while achieving high accuracy (92%), the app misclassified two AD patients as healthy and four healthy controls as having dementia (all classified as AD). Additional details on the classification of healthy controls and dementia patients (AD+DLB) are provided in the supplementary materials.

### App performance

For the dementia group, across all participants, 1,595 valid samples (out of 1,640) were available for analysis in the machine learning algorithm. No imputations were performed for the 45 missing samples. App meta data analysis showed that DLB patients had more missed face responses (U = 122.5, *p = 0.013*) and higher pareidolia scores, though the latter was not significant. Additionally, DLB patients had shorter response times, higher variance, IQR, and coefficient of variation (CV) indicating pronounced response variability when compared to AD group.

Shapley values for the classifier output for gaze tracking showed a trend towards class separation for visual tracking geometry (U = 267.0, p = 0.07), but not for speech (U = 241, p = 0.215). A summary of metrics related to test scores, response times, ocular and speech parameters is shown in Table-2.

**Table-2:** App performance metrics for AD and DLB. Scores for response fluctuations are presented as medians, and for visual perception and motor behavior as means ±standard deviation. Motor behavior outputs from the machine learning model are represented by a.u. – arbitrary units

| **Behavioral Metric** |  | **Alzheimer's disease** | **Dementia with Lewy Body** |
| --- | --- | --- | --- |
| Visual perception | Pareidolias (counts) | $4.57 \pm 5.15$ | $7.65 \pm 9.86$ |
| | Missed response ($*p = 0.013$) | $0.38 \pm 0.67$ | $1.50 \pm 1.85$ |
| | Correct responses | $33.57 \pm 6.57$ | $29.45 \pm 9.95$ |
| Response fluctuation | Per stimulus response time (s) (1<sup>st</sup> quartile – 3<sup>rd</sup> quartile) | 8.01 (5.38 – 9.21) | 6.16 (2.94 – 9.49) |
|  | Coefficient of variation | 0.42 | 0.57 |
|  | Median absolute deviation (robust) | 2.77 | 4.81 |
|  | Variance | 11.24 | 14.79 |
| Motor behavior | Visual tracking geometry (a.u.) | $0.91 \pm 0.40$ | $0.68 \pm 0.49$ |
| | Speech index (a.u.) | $0.13 \pm 0.09$ | $0.11 \pm 0.09$ |
| Probability for a positive classification | | $80.3\% \pm 0.31$ | $83.2\% \pm 0.26$ |

Since raw feature magnitudes differed across variables due to the inherent non-linearity of eye-tracking, CV analysis for Shapley values allowed a degree of explainability for the variability seen between the 2 groups. DLB patients showed a combination of low CV for saccade distances and high CV for number of scans (saccade counts) and clusters (fixation counts) suggestive of short, repetitive saccade amplitudes causing inconsistent visual exploration. However, global fixation duration on targets (faces) remained consistent between the two groups (Figure-3).

**Figure-3:**
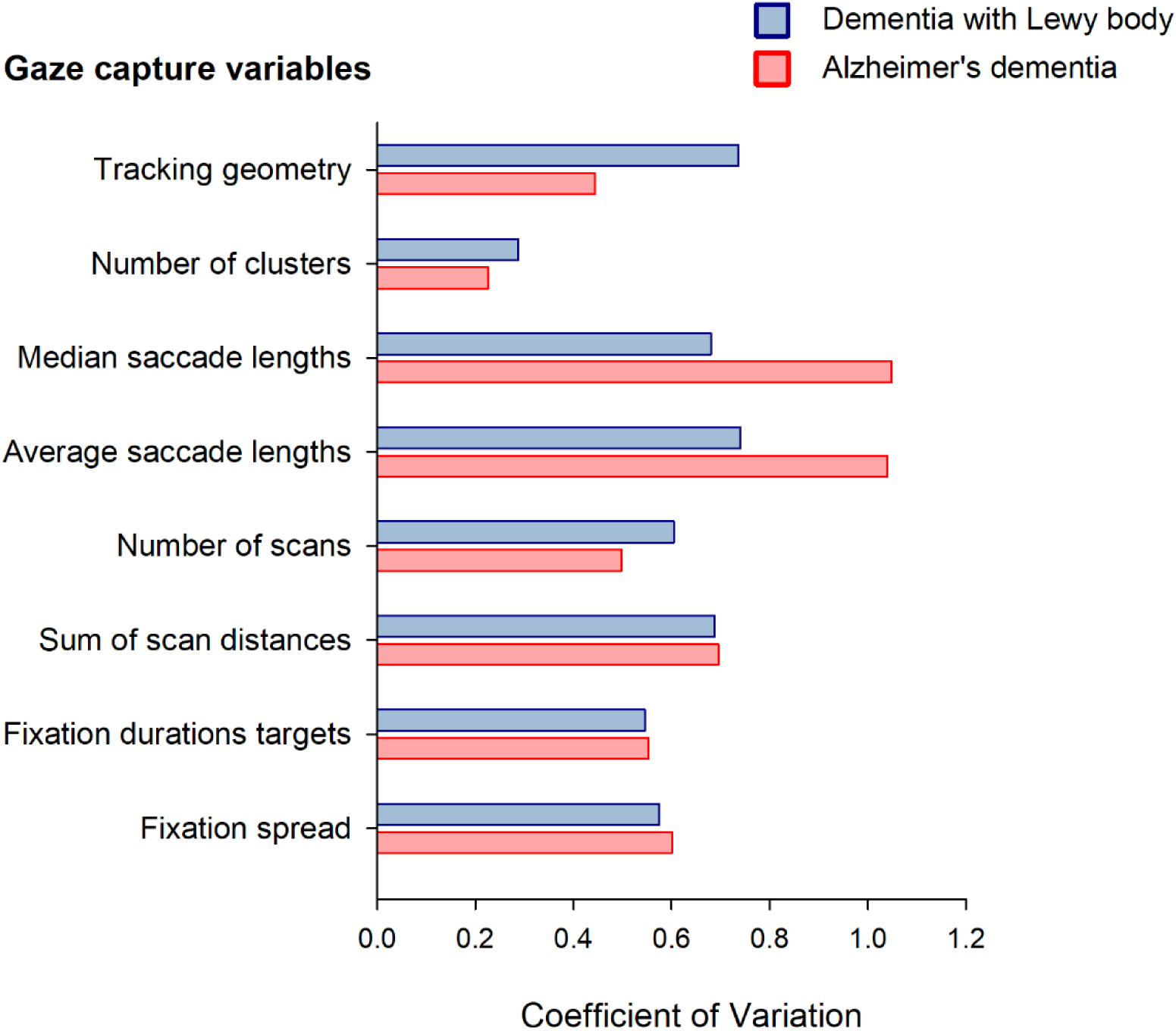
Comparison of coefficient of variation for AD and DLB for different gaze capture features.

Figure-3 shows coefficients of variation for Shapley values generated for gaze capture data. The bar-plot demonstrates a measure of defining the relative stability of feature contribution between AD and DLB. The visual search geometry for DLB in the app test revealed patterns reflecting fluctuating attentional control and impaired visual exploration. To provide additional context, mean and standard deviation values for key features are presented in Supplementary Table-2.

### Diagnostic performance

Table-3 shows the diagnostic performance metrics for AD and DLB when the diagnosis was confirmed via one or more of the following nuclear imaging methods ➔ IMP-SPECT, DAT-SPECT or MIBG. Bayesian binary classification for MMSE-J, paper-based, and digital pareidolia tests showed moderate accuracy (62–65%) with comparable sensitivity (∼60%) and specificity (∼65%). In contrast, the app-based test outperformed all other methods, achieving the highest accuracy (90%) with a sensitivity of 87% and specificity of 93%.

**Table-3:** Diagnostic performance metrics of testing modalities against nuclear imaging diagnosis for classifying AD and DLB patients **Table 3 Legend**: MMSE-J = Mini mental state examination, Japanese version; NPT = Noise pareidolia test; All scores are presented as means ±standard deviation.

| Tests | Optimal Threshold | Sensitivity | Specificity | Positive predictive value | Negative predictive value | Accuracy |
| --- | --- | --- | --- | --- | --- | --- |
| MMSE-J | 21 | $0.57 \pm 0.08$ | $0.65 \pm 0.08$ | $0.53 \pm 0.09$ | $0.69 \pm 0.08$ | $0.62 \pm 0.05$ |
| Paper NPT score | 3 | $0.57 \pm 0.08$ | $0.65 \pm 0.08$ | $0.54 \pm 0.09$ | $0.68 \pm 0.08$ | $0.62 \pm 0.06$ |
| Digital NPT score | 4 | $0.60 \pm 0.08$ | $0.68 \pm 0.08$ | $0.57 \pm 0.09$ | $0.71 \pm 0.07$ | $0.65 \pm 0.06$ |
| App test | 0.6 | **$0.87 \pm 0.06$** | **$0.93 \pm 0.04$** | **$0.90 \pm 0.06$** | **$0.90 \pm 0.05$** | **$0.90 \pm 0.04$** |

ROC analysis indicated that an optimal threshold cutoff of 60% probability provided the highest classification accuracy for the app. Figure-3 shows ROC curve and the corresponding thresholds and probabilistic estimates for the diagnostic performance for the app.

Figure-4A shows Bayesian ROC curve representing the app classifier performance. Blue density contour shows the optimal operating region with the dashed contour line representing the credible intervals at that area. Grey dot defines the optimal threshold (at 60%) for our dataset.

Figure-4B shows the best PPV and NPV (of 90% each) for the optimal threshold.

Figure-4C shows interval plot of probability estimates for key performance metrics with grey circle representing mean and intervals as 95% credible intervals.

**Figure-4:**
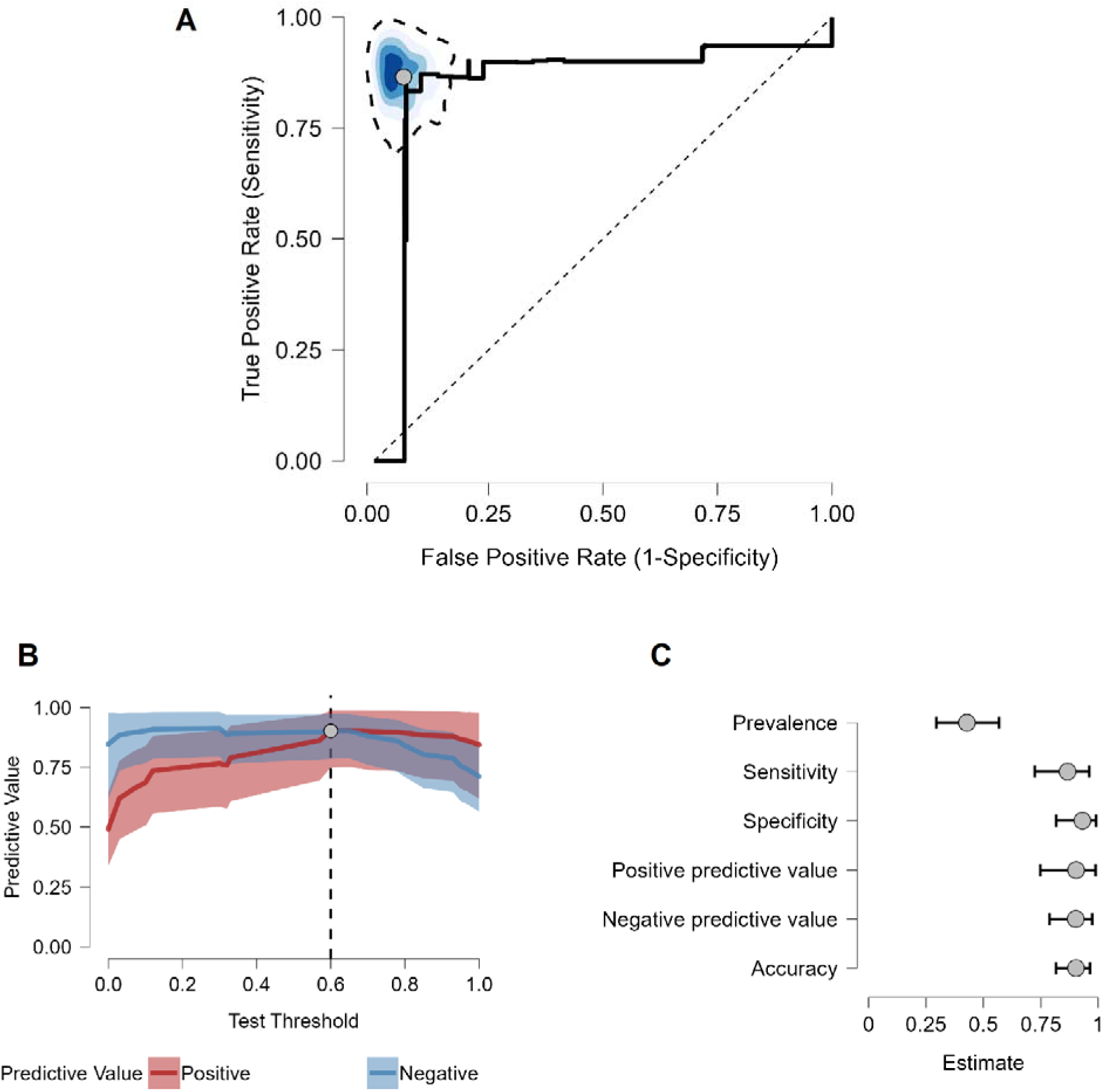
ROC curve and estimates of diagnostic performance for the app.

## Discussion

To our knowledge, this is the first smartphone-based, machine-learning driven study to differentiate dementia with Lewy bodies (DLB) from Alzheimer’s disease (AD) integrating in house developed eye tracking and speech processing algorithms. Our findings demonstrate that (i) conventional neuropsychological assessments such as MMSE and NPT exhibit moderate sensitivity (∼60%) and specificity (∼65%) for DLB differentiation, (ii) core behavioral signatures for DLB were effectively captured using a smartphone-based assessment app platform, and (iii) the app outperformed screening methods with a sensitivity of 87%, specificity of 93%, and overall accuracy of 90% for AD–DLB classification. This ability to identify and quantify subtle behavioral signatures not only emphasizes its potential as a screening tool but also indicates that digital biomarkers may provide a reliable surrogate for gold standard nuclear imaging techniques.

In our dementia cohort, we found that the MMSE and NPT capture important yet overlapping components of cognition. The moderate accuracies of ∼62% in each of these tests could be attributed to their domain specific ability to discriminate neuropsychiatric behavior. MMSE broadly assesses global cognitive domains, but lacks specificity in perceptual and executive dysfunction^44^. In contrast, the NPT directly quantifies visuo-perceptual deficits (pareidolias) that affect non-cognitive domains like visual attention, perceptual integration, and top-down modulation^34^, establishing it as a valuable tool for DLB screening. However in our cohort, inconsistencies in pareidolic behavior in AD, particularly in those without a history of hallucinations, complicated the differentiation of DLB-specific patterns, potentially impacting classification accuracy.

The superiority of the app compared to traditional NPT assessment is largely attributable to its capacity to integrate multiple behavioral dimensions. By design, the NPT is a search-and-detect-paradigm for visuo-perception^21^. Building on this foundation, our approach aimed to define additional core symptoms of DLB, that included parkinsonism features and fluctuating cognition. For instance, integrating eye-tracking metrics from the smartphone front-facing camera provided deeper insights into the visuo-perceptual deficits underlying visual hallucinations^34^. DLB patients within our study population exhibited several fixations and a pattern of multiple, shorter saccades—a pattern consistent with bradykinetic movements seen in parkinsonian syndromes^45^. A combination of reduced variability in saccade distances and increased scanning behavior highlights impaired visual attention in such patients that requires generation of extra saccades to integrate information from noisy stimuli^34^. This may signify disruption in early attention and fixation behavior that is commonly seen in Lewy type of neurodegeneration^46,47^. However, we did not observe any significant differences in fixation durations or fixation time between AD and DLB patients, which is a characteristic of DLB / PDD behavior. We speculate that this could be due to several factors such as overlaps in perceptual processing between AD and DLB^47^, the influence of screen size on eye movement patterns or hardware limitations of the front-facing camera due to limited sampling rate.

Speech analysis is known to act as an early indicator of the spectrum of parkinsonism seen in DLB^48,49^. In our study, speech played a dual role—first, in terms of usability through an intuitive interface, and second, serving as an input for the classification algorithm. The app was designed with minimal touch-based interactions, prioritizing interactive simplicity and a patient-centric user experience. Consistent with prior research employing speech classifier approaches^50^, we obtained approximately two minutes of speech per participant, ensuring sufficient data for analysis. Mapping acoustic spectral properties and response latency provided valuable insights into motor planning and execution efficiency, but these measures did not exhibit sufficient specificity for DLB, thereby complicating the interpretation of the derived speech index. In our current implementation, the visual modality appears to predominantly drive the classifier output relative to the speech component. However, integrating an expanded array of speech features such as vocal intensity, pitch modulation, and linguistic complexity may be pivotal for enhancing overall classifier performance.

Cognitive fluctuations are often challenging to quantify objectively due to their transient and variable nature^11^. We speculated that NPT, designed to evoke illusionary perceptions through ambiguous, sensory-deprived stimuli, may induce cognitive fluctuations by requiring constant target encoding that affects temporal responses related to attention and alertness^37^. Unlike physical examinations, which often miss these transient changes, the app allowed quantification of response variability with high consistency. In our test group, AD patients exhibited more stable response times, with less variability across trials. In contrast, DLB patients had more frequent, irregular, and greater dispersions in stimuli time distributions within the ∼10 min core test time window, demonstrating substantial response fluctuations. On a timescale of seconds to minutes, these results align with prior studies indicating that cognitive fluctuations in DLB are associated with unstable attentional states and impaired perceptual gating^51^.

The integration of machine learning to quantify these microfluctuations in multiple behavioral domains allowed us to subtype AD and DLB with high sensitivity and specificity. Mechanistically, these outputs provided a detailed characterization of non-cognitive symptoms defined via the NPT. These domains implicate a network that includes frontal regions—such as the dorsolateral prefrontal cortex^52^, frontostriatal circuits^34^, and frontal eye fields—essential for integrating top–down modulation with sensory input, and subcortical structures like the basal ganglia, critical for dopaminergic signaling^23^. Specifically the contributions of eye tracking output from our experiments underscores the dysregulation in cholinergic and dopaminergic networks has been shown to correlate with DLB-related behavioral patterns^22^. However, a common challenge in neurodegenerative diagnostics is the ambiguous correlation between nuclear imaging findings (e.g., dopamine degeneration) and observed downstream behavioral dysfunction^53,54^. In this sense, our technology sidesteps this issue by focusing on behavioral outputs, capturing the functional impact of neurodegeneration with a fidelity comparable to nuclear imaging. With complexities in deconstructing neuropsychiatric symptoms, our tool could serve as a valuable adjunct to imaging techniques, providing a more holistic view of disease pathology and progression.

### Implications of this study

DLB is currently the most expensive form of dementia^55,56^. In the absence of disease-modifying treatments, early and accurate classification of DLB is essential not only for ensuring patients receive appropriate care, but also for healthcare systems to optimize resource allocation and reduce healthcare burdens. The app offers a practical, non-invasive, and cost-effective tool that can be easily deployed in various healthcare settings, including resource-limited environments. Given the encouraging outcomes of this study, future studies with diverse cohorts and longitudinal studies could provide valuable insights into how digital biomarkers evolve over the course of the disease, potentially serving as early predictors of progression and treatment response.

### Limitations

Due of the nature of visuo-perceptual testing, we specifically recruited patients who were entirely off antipsychotic medications to eliminate potential drug-induced effects on eye-tracking outcomes^57^. However, along with antipsychotics, the effect of Levodopa and Donepezil medication on visuo-perception should also be carefully studied in the future. The severity of dementia among patients was also not strictly controlled (MCI-level, moderate, or severe type) as the patients were enrolled based on the feasibility of smartphone usage and availability of nuclear imaging results as ground truth. Future optimizations to the model with a larger sample size could enable disease staging, further increasing the clinical relevance of this work. Additionally, testing on more heterogenous populations will be essential in validating the app’s utility and the generalizability of our findings.

It is also important to address the false negative cases in our study. The app misclassified two AD patients as healthy elderly. One was in their 50s, diagnosed a year ago, with a dementia probability score of 15%. The second was in their 70s, also diagnosed an year ago with a probability of 45%. While the dementia spectrum is inherently heterogeneous, these misclassifications suggest potential gaps in the model’s sensitivity to atypical disease presentations, such as early-onset cases with preserved cognitive reserves or subtle phenotypic variations such as mixed dementia cases. Our digital approach captures key behavioral dimensions but may not fully account for symptom heterogeneity across neurodegenerative disorders. Further refinement of the app’s algorithms is needed to enhance its ability to distinguish dementia subtypes beyond AD and DLB.

### Conclusions

Our study demonstrates that the app, through digital profiling of behavior and the integration of machine learning algorithms, offers a precise and efficient method for differentiating between DLB and AD compared to traditional behavioral assessments. Its performance aligns closely with that of established nuclear imaging modalities, underscoring its potential as a valuable tool for clinical use. By effectively capturing core symptoms, our smartphone-based modality provides a reliable, scalable, and non-invasive tool for classification of DLB from AD. Future studies that incorporate multimodal digital assessments are essential to further enhance diagnostic precision and to broaden our understanding of neurodegenerative disease processes.

## Data Availability

All data produced in the present study are available upon reasonable request to the authors

## Acknowledgements

Our sincere thanks to the individuals who participated in the study; to the directors and clinical staff of Asakayama hospital, Nippon life Hospital and Osaka University hospital for their assistance in the data collection.

## Author contributions

Conceptualization: GSR, EM.

Methodology: GSR, MS, HK, YN, SSR, HM, MI, EM.

Software: GSR, ACS, SSR.

Investigation: GSR, TO, MS, HK, KS, YN, KY, YY, IO, NY, CS, DN, KF, HM, EM, MI, KN.

Validation: GSR, ACS, EM.

Formal analysis: GSR, ACS, AW.

Resources: GSR, EM, SSR, HM, MI, KN.

Data Curation: GSR, TO, ACS.

Writing - Original Draft: GSR

Writing - Review & Editing: GSR, TO, ACS, MS, HK, KS, YN, KY, YK, YY, SSR, AW, MH, YN, HM, EM, MI, KN.

Visualization: GSR, ACS, EM

Supervision: GSR, DN, EM, MI, HM, KN

Project administration: GSR

Funding acquisition: GSR

## Declaration of generative AI and AI-assisted technologies in the writing process

During the preparation of this work the author(s) used [ChatGPT] in order to [restructure sentences]. After using this tool/service, the author(s) reviewed and edited the content as needed and take(s) full responsibility for the content of the publication.

