## Supplementary Data for "Smartphone-based behavioral profiling for distinguishing Dementia with Lewy bodies from Alzheimer’s Disease"

Revankar et.al.

**Supplementary information**

**Supplementary Table-1: App performance metrics for Healthy controls and Dementia**

| **Behavioral Metric** | | **Healthy controls (N=40)** | **Dementia  (N=41)** | **p-value** |
| --- | --- | --- | --- | --- |
| Core test completion time (min) | | 5.51 ± 1.73 | 8.17 ± 2.71 | <0.001 |
| Response fluctuation | Per stimulus response time (s) (1^st^ quartile – 3^rd^ quartile) | 5.04 (3.51 – 5.94) | 7.80  (3.92 – 9.21) | 0.004 |
|  | Coefficient of variation | 0.41 | 0.47 | - |
|  | Median absolute deviation (robust) | 1.59 | 3.02 | - |
|  | Variance | 10.21 | 17.27 | - |
|  | Interquartile range | 2.38 | 4.25 | - |
| Visual perception | Pareidolias (counts) | 1.17 ± 2.55 | 6.07 ± 7.87 | <0.001 |
|  | Missed response | 0.10 ± 0.30 | 0.92 ± 1.47 | <0.001 |
|  | Correct responses | 38.40 ± 2.83 | 31.56 ± 8.55 | <0.001 |
| Motor behavior | Visual tracking geometry (a.u.) | 0.87 ± 0.54 | 0.79 ± 0.46 | 0.260 |
|  | Speech index (a.u.) | 0.09 ± 0.08 | 0.12 ± 0.09 | 0.068 |
| Probability for a positive classification | | 80.4% ± 0.21 | 90.0% ± 0.17 |  |

Supplementary Table 1 Legend: a.u. – arbitrary unit; Scores for response fluctuations are presented as medians and for visual perception and motor behavior as means ±standard deviation

**Supplementary Table-2: Diagnostic performance metrics of testing modalities against clinical examination of healthy and dementia participants**

| **Tests** | **Optimal Threshold** | **Sensitivity** | **Specificity** | **Positive predictive value** | **Negative predictive value** | **Accuracy** |
| --- | --- | --- | --- | --- | --- | --- |
| MMSE-J | 27 | 0.90 ± 0.04 | 0.90 ± 0.04 | 0.88 ± 0.05 | 0.92 ± 0.04 | 0.90 ± 0.03 |
| Digital NPT score | 2 | 0.84 ± 0.05 | 0.74 ± 0.06 | 0.73 ± 0.06 | 0.85 ± 0.05 | 0.79 ± 0.04 |
| App test | 0.35 | **0.94 ± 0.03** | **0.90 ± 0.04** | **0.89 ± 0.05** | **0.95 ± 0.03** | **0.92 ± 0.03** |

Supplementary Table-2 Legend: MMSE-J = Mini mental state examination, Japanese version; NPT = Noise pareidolia test; All scores are presented as means ±standard deviation.

**Supplementary Figure-1: ROC and estimates for Healthy controls vs Dementia**

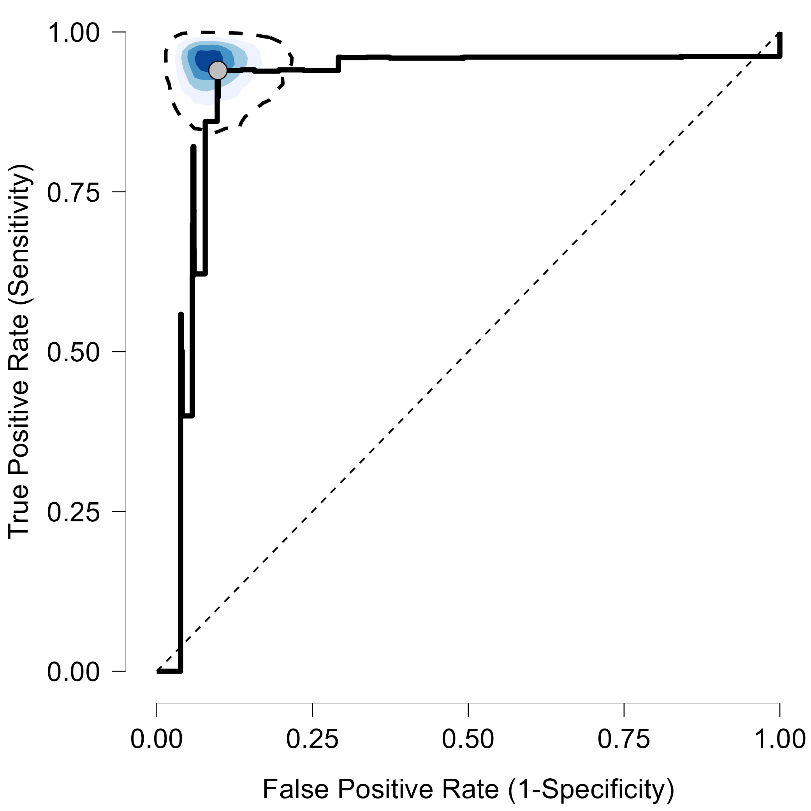

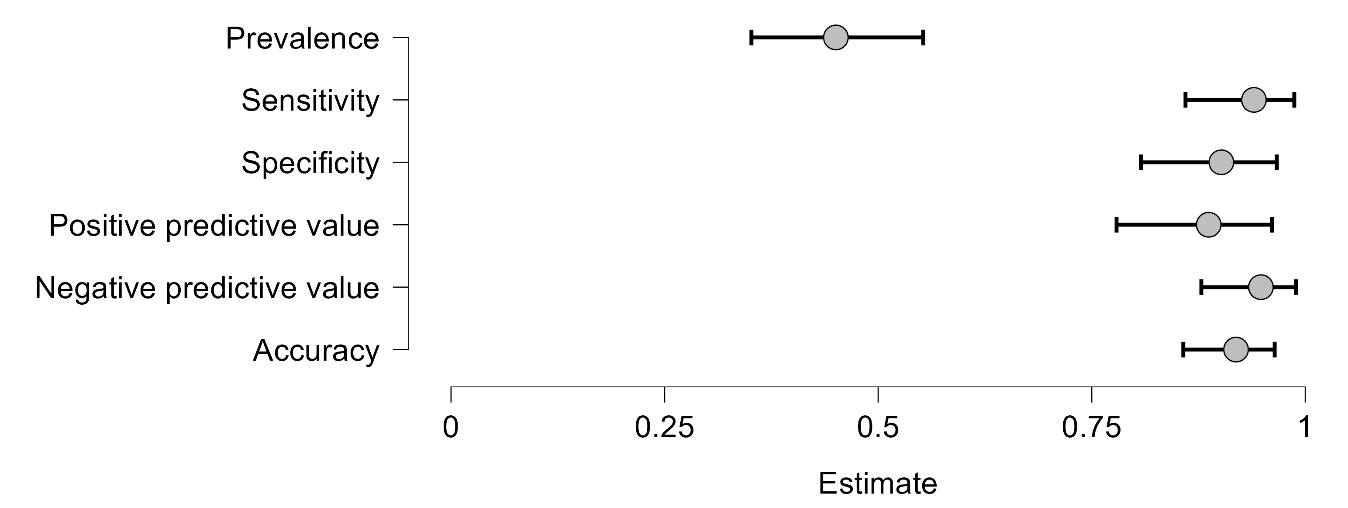

Supplementary Figure-1 (top) shows Bayesian ROC curve representing the app classifier performance. Blue density contour shows the optimal operating region with the dashed contour line representing the credible intervals at that area. Grey dot defines the optimal threshold at 35%. Figure (bottom) shows interval plot of probability estimates for key performance metrics with grey circle representing mean and intervals as 95% credible intervals.

**Supplementary Table-3: Mean and SDs of Shapley values for probability scores generated for Alzheimer’s disease and dementia with Lewy body.**

| **Metric** | **Alzheimer’s disease** | **Dementia with Lewy body** | **p-value** |
| --- | --- | --- | --- |
| Tracking geometry | 0.90 ± 0.40 | 0.67 ± 0.49 | 0.071 |
| Number of Clusters | 0.002 ± 0.00 | 0.002 ± 0.00 | 0.220 |
| Median Saccade Distances | 0.04 ± 0.04 | 0.02 ± 0.01 | 0.081 |
| Average Saccade Distances | 0.015 ± 0.02 | 0.008 ± 0.01 | 0.074 |
| Number of Scans | 0.49 ± 0.24 | 0.33 ± 0.19 | 0.028 |
| Sum of scan distances | 1.47 ± 1.02 | 0.80 ± 0.55 | 0.007 |
| Fixation duration, targets | 0.017 ± 0.01 | 0.014 ± 0.01 | 0.351 |
| Fixation spread | 0.076 ± 0.05 | 0.078 ± 0.04 | 0.901 |

Supplementary Table-3 shows mean, SDs of Shapley values for gaze parameters that contributed to the probability values for the classifier (at the second stage). Since raw data from the variables were non-linear, the representation of coefficient of variation was a better measure for interpretation.
